# History of incarceration and age-related cognitive impairment: Testing models of genetic and environmental risk in longitudinal panel study of older adults

**DOI:** 10.1101/2023.06.26.23291910

**Authors:** Peter T. Tanksley, Matthew W. Logan, J.C. Barnes

## Abstract

History of incarceration is associated with an excess of morbidity and mortality. While the incarceration experience itself comes with substantive health risks (e.g., injury, psychological stress, exposure to infectious disease), most inmates eventually return to the general population where they will be diagnosed with the same age-related conditions that drive mortality in the non-incarcerated population but at exaggerated rates. However, the interplay between history of incarceration as a risk factor and more traditional risk factors for age-related diseases (e.g., genetic risk factors) has not been studied. Here, we focus on cognitive impairment, a hallmark of neurodegenerative conditions like Alzheimer’s disease, as an age-related state that may be uniquely impacted by the confluence of environmental stressors (e.g., incarceration) and genetic risk factors. Using data from the Health and Retirement Study, we found that incarceration and *APOE-ε4* genotype (i.e., the chief genetic risk factor for Alzheimer’s disease) both constituted substantive risk factors for cognitive impairment in terms of overall risk and earlier onset. The observed effects were mutually independent, however, suggesting that the risk conveyed by incarceration and *APOE-ε4* genotype operate across different risk pathways. Our results have implications for the study of criminal justice contact as a public health risk factor for age-related, neurodegenerative conditions.

## Introduction

The effects of incarceration are insidious, pervasive, and correspond with a litany of physical and mental health afflictions among offenders.(1–7) Although the causal mechanisms underlying these relationships are nuanced, there is considerable evidence to suggest that imprisonment exacerbates pre-existing medical conditions and other risk factors due to high rates of exposure to environmental stressors, such as prison violence and victimization, and low rates of identification and treatment during the intake process. Research also indicates that the effects of incarceration are durable and extend into the communities to which individuals return, negatively impacting their quality of life and expectancy.(8)

Of particular importance to practitioners and policymakers is the influence of the incarceration experience on acute and chronic causes of early mortality. In the time period around their release, people leaving prison are more likely to die from opioid use disorders (OUDs), overdose, suicide, and homicide.(9–17) One large-scale review on incarceration and health found that most post-release mortality studies focus on short-term effects and indicate that the period immediately following release represented a high-risk time for early mortality among former inmates due to acute causes.(18)

The long-term impact of incarceration on mortality (i.e., via chronic health conditions) is less well understood. Most former inmates survive the high-mortality period immediately following release and eventually succumb to the same chronic conditions that drive mortality for the general population such as cancer, cardiovascular disease, and neurodegenerative diseases such as Alzheimer’s Disease and Related Dementias (ADRDs).(19–23) Our focus here is on the latter affliction; specifically, the potential long-term effects of incarceration on the onset and progression of neurodegeneration as indicated by cognitive impairment.

Relative to other chronic conditions, neurodegenerative diseases and their potential link with incarceration history have received less attention(24) (but see (25–27)). Still, existing research generally finds that age-related cognitive impairments (i.e., the chief indicator of neurodegeneration) are more prevalent among those who have been incarcerated at some point in their lives compared to the general population.(19, 20, 22) This is unsurprising, as former inmates are subject to a constellation of risk factors including acute stressors (e.g., traumatic brain injury, high infectious disease burden), chronic stressors (e.g., disenfranchisement, unemployment), and lack of social integration (e.g., difficulty maintaining familial connections, homelessness).(18, 28) Together, incarceration and its collateral consequences have been identified as powerful environmental risk factors for the development and progression of neurodegenerative conditions. However, it is not well-understood if/how the risks associated with the incarceration experience interact with the other main source of risk for neurodegeneration: genetic risk factors.

Neurodegenerative diseases possess complex genetic etiologies. Alzheimer’s disease (AD), for example, is oligogenic, meaning it is primarily influenced by a small number of genetic variants. For instance, apolipoprotein E (*APOE*) is a gene that codes for proteins that bind and transport low-density lipids and contribute to cholesterol clearance from the bloodstream.(29) Variation in this gene can impact cholesterol metabolism and may lead to increased risk for stroke, cardiovascular disease, and diagnosis of AD.(30) The *APOE* genotype is determined by two variants (rs7412 and rs429358), resulting in three main isoforms of protein *apoE*: E2, E3, and E4 encoded by the ε2, ε3, and ε4 alleles, respectively. The *APOE-ε4* allele has been linked with substantial increases in the risk of developing late-onset Alzheimer’s disease. Possession of one copy of the *APOE-ε4* allele confers a 3-fold increase in risk while two copies confer a 15-fold increase, making *APOE-ε4* status the strongest genetic predictor of Alzheimer’s disease risk.(31) Recent genome-wide association studies have begun to illuminate polygenic variation that contributes to AD(32), but highly impactful genes like *APOE* still loom large in the landscape of genetic risk.

Despite their strong genetic basis, neurodegenerative diseases are also affected by environmental risk factors. The 2020 Lancet Commission report on dementia prevention, intervention, and care identified 12 modifiable risk factors that, together, accounted for 40% of dementias world-wide.(33) These included education, hearing loss, traumatic brain injury, hypertension, alcohol intake, obesity, smoking, depression, social isolation, physical inactivity, diabetes, and air pollution. Minoritized groups, particularly African Americans in the US, also bear increased risk for neurodegeneration.(34) Considering these environmental risk factors and overrepresentation of minoritized groups in the correctional system, it is unsurprising that recent research has found that neurodegenerative diseases are more prevalent in the current/former members of the carceral population than among the general population.(35)

Here, we draw these two lines of research together to examine how incarceration history and *APOE-ε4* genotype combine to produce risk for neurodegeneration as indicated by cognitive impairment. We posit four possible scenarios (**Figure 1**): (1) genetic risks dominate prediction of cognitive impairment (“genetic risk model”); (2) environmental risks dominate prediction of cognitive impairment (“environmental risk model”), (3) genetic and environmental risks independently predict cognitive impairment (“G+E model”), and (4) genetic and environmental risks interact to produce multiplicative risk for cognitive impairment (“G×E model”). We test for evidence of these four scenarios in terms of both overall risk (i.e., ever developing cognitive impairment) and progression of cognitive impairment (i.e., age of onset).

**Figure 1.**
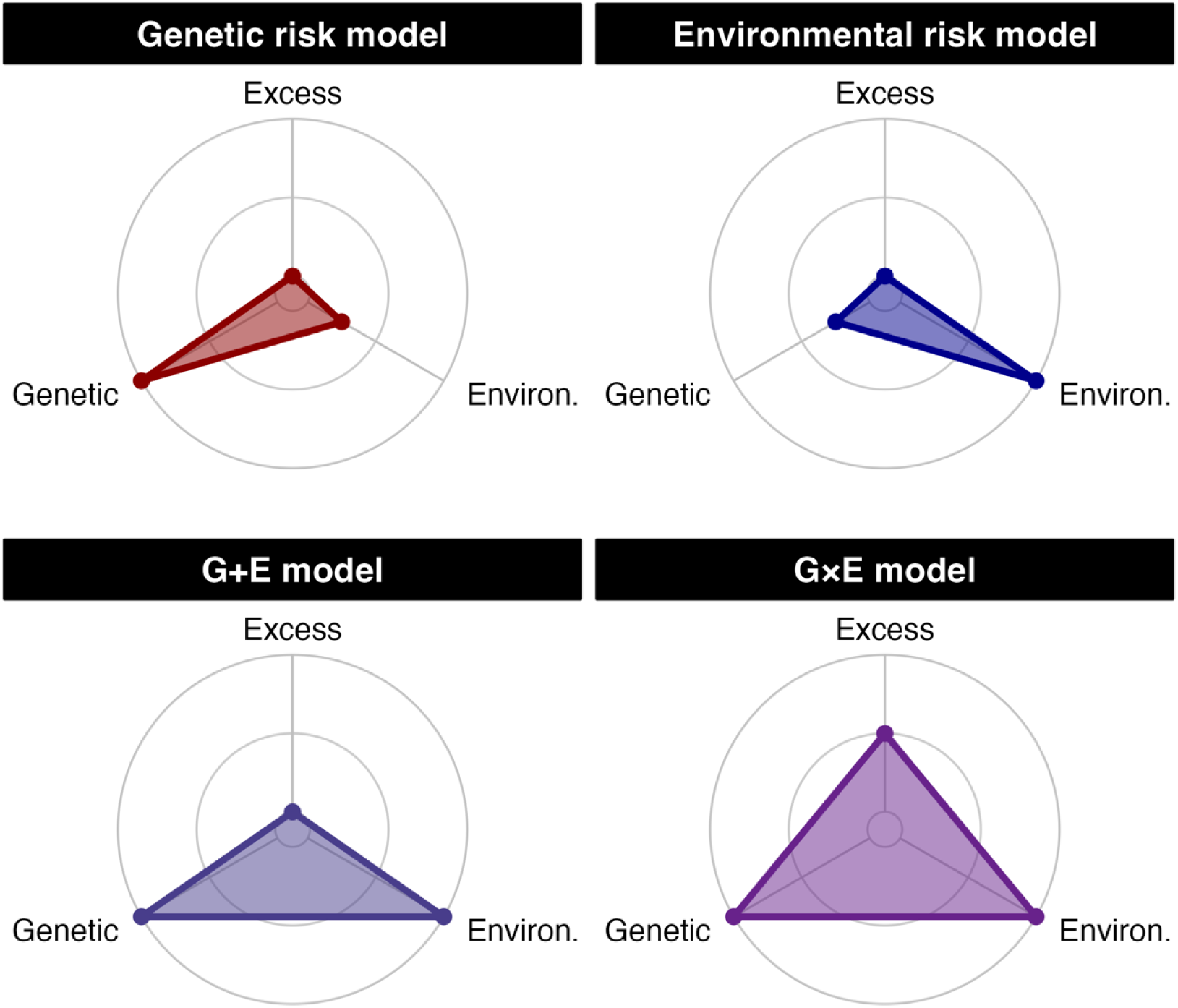
Radar plots showing the theoretical models of prediction sources (genetic, environmental, excess) explored in the current study. Distance from the origin of each plot represents the increasing magnitude of each risk factor’s predictive power. Note: “Excess” refers to the additional prediction achieved by a model containing an interaction term.

## Methods

### Data

Data come from the Health and Retirement Study (HRS), a nationally representative cohort study of elderly Americans aged ≥50.(36) The HRS collects information from participants on a biennial basis on social, demographic, economic, behavioral, and health conditions. In addition to the main survey, the HRS randomly selected half the sample to receive enhanced face-to-face interviews starting in 2006 and alternating to the other half of the sample with each wave of data collection. Enhanced interviews included physical performance measurements, blood and saliva samples, and a leave-behind psychosocial questionnaire. The current study relies on deidentified participant data on cognitive impairment from the main study, as well as *APOE-ε4* genotype and information on lifetime incarceration drawn from the enhanced interviews. The analytic sample with complete data included *N*=6,949 cases with *N*=55,345 observations (see **Figure S1** information on missingness).

### Measures

#### Cognitive impairment

We assessed cognitive status of HRS participants using the classification scheme developed for the HRS. All participants were administered the Telephone Interview for Cognitive Status (TICS), a telephone-based instrument that probed cognitive functioning along seven domains: memory, mental status, abstract reasoning, fluid reasoning, vocabulary, dementia, and numeracy. In 2009, a methodology was developed for using TICS scores to classify respondents as “normal”, “cognitively impaired non-dementia”, and “demented”. The classification system was validated by a team of dementia experts by comparing TICS scores to the Aging, Demographic, and Memory Study (ADAMS), a subsample of the HRS who received a more extensive psychological battery.(37) For the current analysis, we used repeated measures of the Langa-Weir Cognitive Status classifications for assessment years 1998-2020 from the HRS user contributed data set. To avoid small cell counts, we combined both non-normal cognitive states into one “impaired” category, resulting in a dichotomous *cognitive impairment* variable.

#### Lifetime Incarceration

HRS participants were asked about adverse experiences in adulthood including “Have you ever been an inmate in a jail, prison, juvenile detention center, or other correctional facility?”. Respondents received a score of 1 if they answered “Yes” and a score of 0 if they answered “No”. This question was asked in 2012/2014 and answers were coded so that if an individual answered “Yes” to either wave they received a score of 1 on the *Lifetime Incarceration* measure. Cases were only categorized as “missing” if they did not provide responses on both waves of data collection. The resulting variable indicated that approximately 10% of the analytic samples had some history of incarceration (**Table 1**). These observed rates of incarceration align with other national estimates.(38)

**Table 1.** Individual and observation-level descriptive statistics of the Health and Retirement Study sample.

| Variable | Overall, N = 6,949<sup>1</sup> | Ever incarcerated? |  | p-value<sup>2</sup> |
| --- | --- | --- | --- | --- |
|  |  | No, N = 6,244<sup>1</sup> | Yes, N = 705<sup>1</sup> |  |
| **Individuals (N = 6,949)** |  |  |  |  |
| HRS cohort |  |  |  | **<0.001** |
| AHEAD | 511 (7.4%) | 491 (7.9%) | 20 (2.8%) |  |
| CODA | 656 (9.4%) | 638 (10%) | 18 (2.6%) |  |
| WB | 1,318 (19%) | 1,226 (20%) | 92 (13%) |  |
| EBB | 2,289 (33%) | 2,008 (32%) | 281 (40%) |  |
| MBB | 2,175 (31%) | 1,881 (30%) | 294 (42%) |  |
| Self-reported sex |  |  |  | **<0.001** |
| Female | 4,099 (59%) | 3,912 (63%) | 187 (27%) |  |
| Male | 2,850 (41%) | 2,332 (37%) | 518 (73%) |  |
| Race/Ethnicity |  |  |  | **<0.001** |
| White | 4,592 (66%) | 4,243 (68%) | 349 (50%) |  |
| Black | 1,229 (18%) | 996 (16%) | 233 (33%) |  |
| Hispanic | 903 (13%) | 799 (13%) | 104 (15%) |  |
| Other | 225 (3.2%) | 206 (3.3%) | 19 (2.7%) |  |
| Years in study | 14.2 (5.2) | 14.4 (5.2) | 12.0 (4.8) | **<0.001** |
| HS completion |  |  |  | **<0.001** |
| High school or more | 5,880 (85%) | 5,361 (86%) | 519 (74%) |  |
| Less than high school | 1,069 (15%) | 883 (14%) | 186 (26%) |  |
| Ever smoker |  |  |  | **<0.001** |
| Yes | 3,723 (54%) | 3,164 (51%) | 559 (79%) |  |
| No | 3,226 (46%) | 3,080 (49%) | 146 (21%) |  |
| APOE-ε4 count |  |  |  | 0.088 |
| Zero copies | 5,202 (75%) | 4,697 (75%) | 505 (72%) |  |
| One copy | 1,603 (23%) | 1,417 (23%) | 186 (26%) |  |
| Two copies | 144 (2.1%) | 130 (2.1%) | 14 (2.0%) |  |
| Social origins index | 0.91 (1.17) | 0.87 (1.14) | 1.26 (1.33) | **<0.001** |
| **Observations (N = 55,345)** |  |  |  |  |
| Age (mean)<sup>3</sup> | 64.63 (9.83) | 64.94 (9.96) | 61.43 (7.62) | **<0.001** |
| Cognitive function |  |  |  | **<0.001** |
| Impaired | 8,275 (15%) | 7,199 (14%) | 1,076 (22%) |  |
| Normal | 47,070 (85%) | 43,288 (86%) | 3,782 (78%) |  |
| History of stroke |  |  |  | 0.228 |
| Yes | 3,329 (6.0%) | 2,962 (5.9%) | 367 (7.6%) |  |
| No | 52,016 (94%) | 47,525 (94%) | 4,491 (92%) |  |
| Study year |  |  |  | **<0.001** |
| 1998 | 2,278 (4.1%) | 2,167 (4.3%) | 111 (2.3%) |  |
| 2000 | 2,287 (4.1%) | 2,178 (4.3%) | 109 (2.2%) |  |
| 2002 | 2,345 (4.2%) | 2,230 (4.4%) | 115 (2.4%) |  |
| 2004 | 3,971 (7.2%) | 3,678 (7.3%) | 293 (6.0%) |  |
| 2006 | 3,982 (7.2%) | 3,696 (7.3%) | 286 (5.9%) |  |
| 2008 | 4,020 (7.3%) | 3,726 (7.4%) | 294 (6.1%) |  |
| 2010 | 6,812 (12%) | 6,127 (12%) | 685 (14%) |  |
| 2012 | 6,886 (12%) | 6,187 (12%) | 699 (14%) |  |
| 2014 | 6,702 (12%) | 6,022 (12%) | 680 (14%) |  |
| 2016 | 6,024 (11%) | 5,420 (11%) | 604 (12%) |  |
| 2018 | 5,183 (9.4%) | 4,667 (9.2%) | 516 (11%) |  |
| 2020 | 4,855 (8.8%) | 4,389 (8.7%) | 466 (9.6%) |  |

<sup>1</sup> n (%); Mean (SD)
<sup>2</sup> Pearson's Chi-squared test (categorical); Wilcoxon rank sum test (continuous); repeated measures logistic regression with random intercept at the individual level (cognitive function, history of stroke).
<sup>3</sup> Age was averaged across observations and tested for differences across incarceration groups using the Wilcoxon rank sum test.

#### APOE-ε4 genotype

We assess monogenic risk for neurodegeneration by classifying HRS participants according to the number of *APOE-ε4* alleles they carry as the ε4 allele has been linked with large increases in risk for stroke and heart disease and is the leading genetic risk factor for Alzheimer’s disease. *APOE* genotype was assessed in the HRS using two methods. The primary method relied on direct genotyping using TaqMan allelic discrimination SNP assays and saliva samples collected from consenting HRS participants during years 2006, 2008, 2010, and 2012.(39) Among individuals for whom direct genotyping was not conducted, *APOE* genotype was classified using imputed genotype array data. Following Ware and colleagues(40), we removed samples with imputed *APOE* genotypes with INFO Scores of <99%. We coded *APOE-ε4* genotype according to the number of ε4 alleles contained in each participants’ genotype, ranging from 0-2, with more alleles representing greater risk.

#### Covariates

Each model was adjusted for age, self-reported race/ethnicity, self-reported sex, history of ever smoking, history of stroke (ever had/doctor suspected stroke=1), high school completion, social origins index (i.e., childhood financial hardship)(41), and HRS birth cohort.

### Analytic strategy

We estimated the overall risk for cognitive impairment in the HRS as a function of lifetime incarceration and *APOE-ε4* genotype using a generalized mixed linear modeling framework. We estimated incidence rate ratios (IRRs) of cognitive impairment using Poisson regression models with a random intercept at the individual level to account for non-independence due to multiple observations. This approach removes interrelatedness among observations from the same individual, allowing for the interpretation of results as if they were drawn from a cross-sectional sample of unrelated individuals. Next, we estimated the hazard for onset of cognitive impairment using two time-to-event methods. First, using nonparametric Kaplan-Meier survival curves, we compared the unadjusted differences between strata of interest (e.g., incarcerated vs. never-incarcerated; *APOE-ε4* allele counts of zero vs. one. vs. two) using log-rank tests. Next, covariate adjustment was accomplished using Cox proportional hazard models. Time-varying covariates (e.g., stroke) were accommodated by creating panel datasets with one row per individual per observation. Both survival methods used chronological age as the time-scale, an alternative to the time-on-study approach, because it is recommended for analyses in elderly cohorts with age-dependent outcomes.(42, 43) According to recommendations for age-scale survival models(44), we addressed possible calendar effects by stratifying Cox models by HRS cohort (i.e., allowing each birth cohort to have a different baseline risk). Additionally, as the age-based time-scale effectively accounts for age, we did not include age as a covariate in the Cox models. The proportionality assumption was confirmed for all Cox models by examining Shoenfeld residuals.

## Results

### Do genetic risk factors predict cognitive impairment in older adults?

We found that *APOE-ε4 genotype* was associated with an increased risk of impaired cognitive function during the study period (**Table 2**). Consistent with prior research, we observed that, relative to those with no *APOE-ε4* alleles, risk for cognitive impairment increased as *APOE-ε4* allele count increased from one allele (IRR=1.12, 95% CI [1.01-1.23], P=0.025) to two (IRR=1.48, 95% CI [1.12-1.95], P=0.006) (**Model 2.1**). When estimated in the same model as *Lifetime incarceration* (**Model 2.3**) the results were essentially unchanged, suggesting that the monogenic risk for cognitive impairment conferred by *APOE-ε4 genotype* is independent of *Lifetime incarceration*.

**Table 2.**
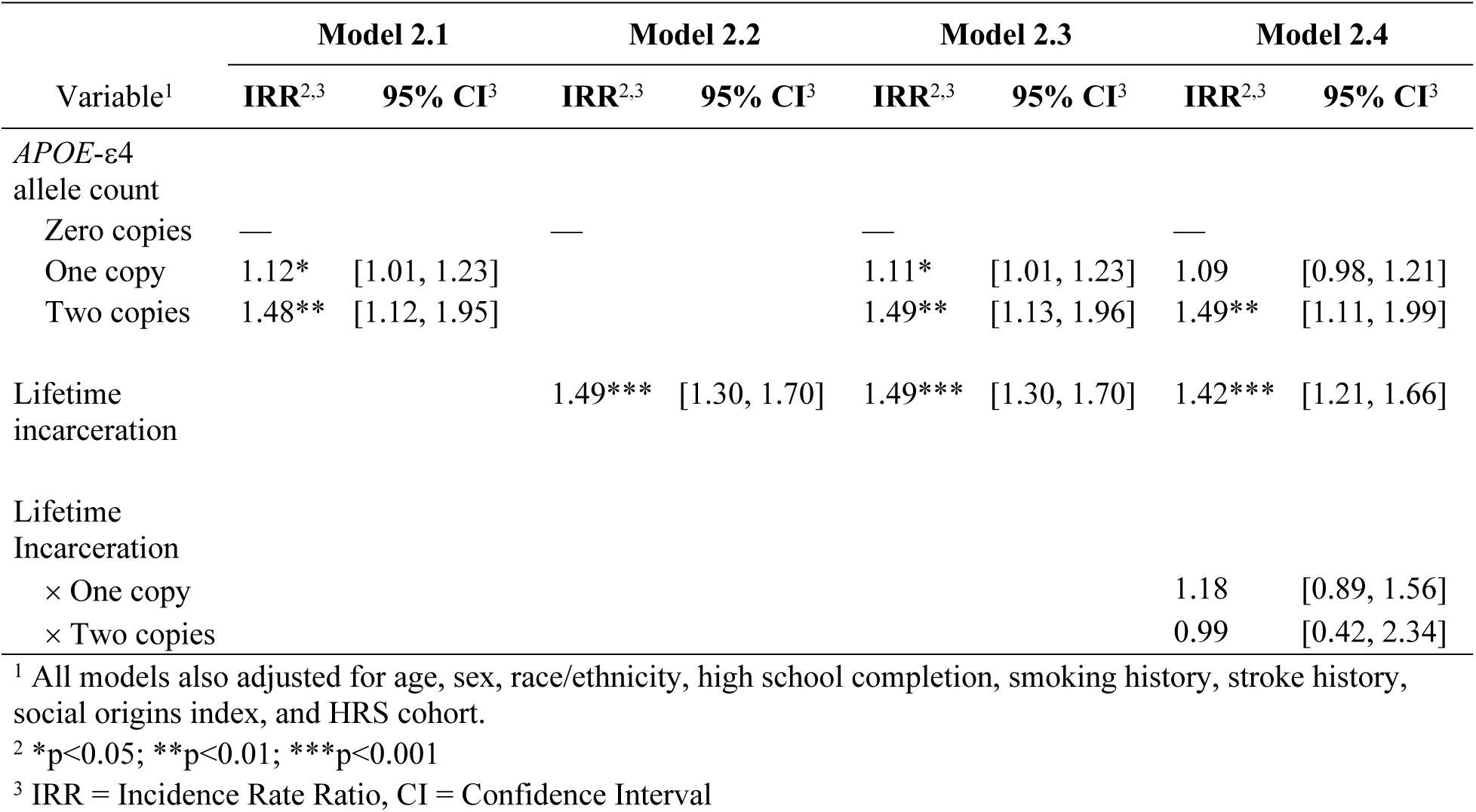
Mixed effect Poisson regression of cognitive status on *APOE*-ε4 genotype and lifetime incarceration

### Is lifetime incarceration a risk factor for cognitive impairment in older adults?

We found that *Lifetime incarceration* conferred a substantive increase in the risk of cognitive impairment (**Table 2**). HRS participants with a history of incarceration were 1.5 times as likely to be cognitively impaired than their non-incarcerated peers (IRR=1.49, 95% CI=[1.30, 1.70], P<0.001) (**Model 2.2**)–a level of risk equal to that for individuals with two *APOE-ε4* alleles. As before, we found that association between *Lifetime Incarceration* and *Cognitive Impairment* were virtually unchanged when *APOE-ε4 genotype* was included in the model (**Model 2.3**) suggesting independent influences on impairment in the sample.

These findings support the interpretation that *APOE-ε4 genotype* and *Lifetime incarceration* operate as independent risk factors for *Cognitive impairment* in later adulthood. The possibility remains, however, that these genetic and environmental sources of risk, when brought together, will go beyond their additive effects to inflict a multiplicative increase in risk for impairment. We test this possibility in the next section.

### Does the incarceration experience amplify genetic risk factors for cognitive impairment in older adults?

We did not find evidence that *Lifetime incarceration* interacts with *APOE-ε4 genotype* to produce excess risk for cognitive impairment. We entered multiplicative interaction terms for *Lifetime incarceration* and *APOE-ε4 genotype* into our analysis (**Model 2.4**) and found that neither term predicted excess risk of cognitive impairment that was distinguishable from the additive effects. Re-estimating **Model 2.4** as a linear model under a linear probability framework produced the same null results, suggesting the lack of interaction was not due to the scale of the interaction (i.e., multiplicative vs. additive).(45) This finding supports the interpretation that *Lifetime incarceration* and *APOE-ε4 genotype* convey their risk for later life cognitive impairment in a fashion that is independent and additive in nature.

### Do genetic risk factors for AD predict earlier cognitive impairment?

In addition to conferring more risk for cognitive impairment at any point (reported above), we found that *APOE-ε4 genotype* was associated with risk for earlier onset as well. We estimated univariate Kaplan-Meier survival curves using age at first indication of cognitive impairment and found statistically significant differences across *APOE-ε4 genotype* according to a log-rank test (**χ**^2^(2)=45.3, P<0.001) (**Figure 2A**). The ages of median survival probability across *APOE-ε4* genotypes were 79 (zero copies of *APOE-ε4*), 74 (one copy of *APOE-ε4*), and 72 (two copies of *APOE-ε4*). We adjusted for covariates with a Cox proportional hazard model using panel datasets to accommodate time-dependent covariates (e.g., stroke history). We observed that possession of one (HR=1.19, 95% CI [1.08-1.31], P<0.001) or two copies (HR=1.56, 95% CI [1.20-2.02], P<0.001) of *APOE-ε4* resulted in significantly greater hazard for developing cognitive impairment in later life (**Table S1, Model S1.1**).

**Figure 2.**
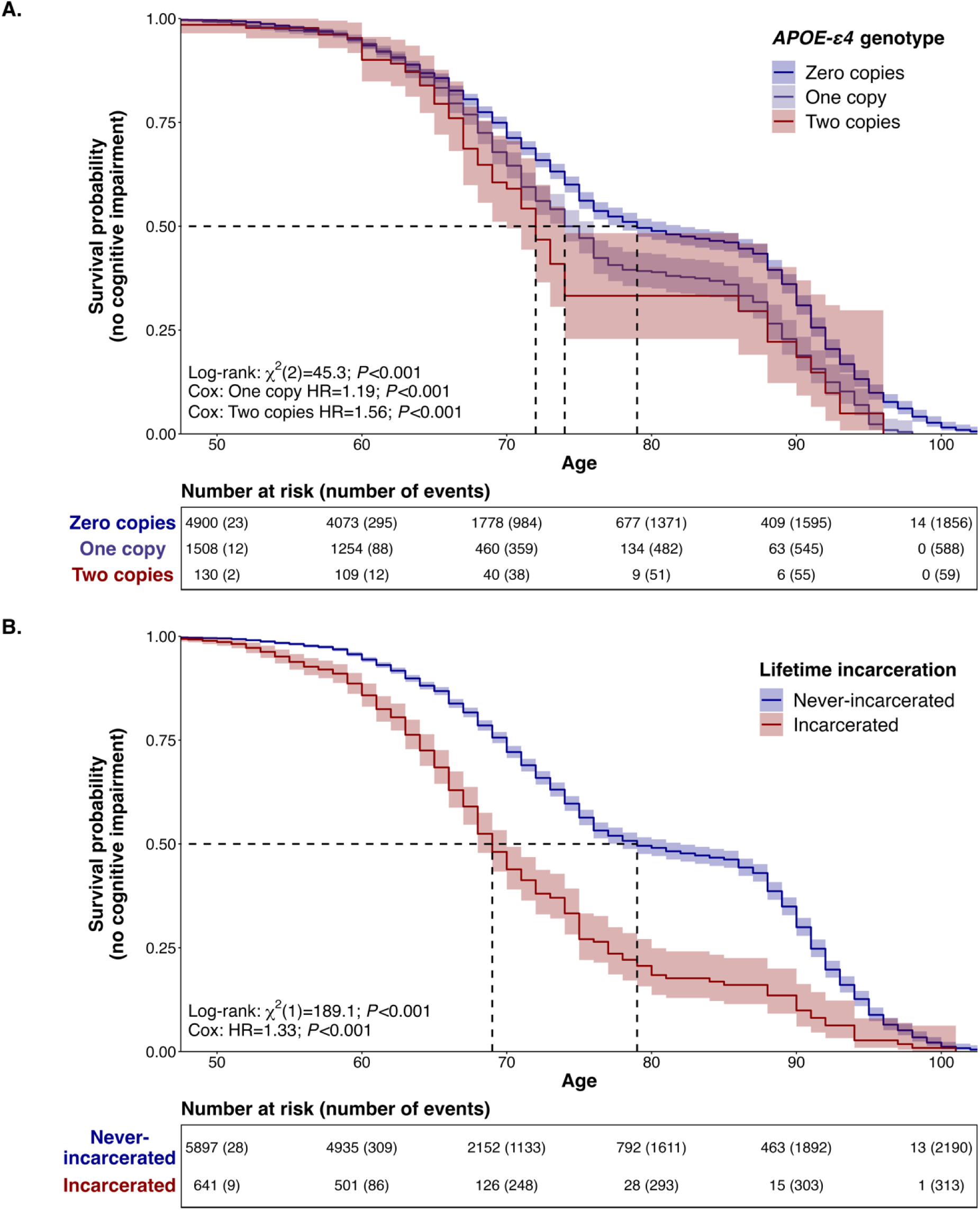
Non-parametric Kaplan-Meier survival curves and risk tables for onset of cognitive impairment, stratified by *APOE-ε4* genotype (**Panel A**) and lifetime incarceration (**Panel B**), respectively. Time-scale corresponds to chronological age. Dashed lines indicate age at median survival probability per stratum. Number of events (i.e., first designation of cognitive impairment) are cumulative. Note: individuals were considered censored for any non-event status (e.g., death, non-participation); x-axis used a floor of age 50 for clarity.

### Does lifetime incarceration predict earlier cognitive impairment?

We found evidence that HRS participants with a history of incarceration were at risk for earlier cognitive impairment compared to those without. Comparing Kaplan-Meier survival curves, we found that individuals with a history of incarceration tended to develop cognitive impairment earlier than their never-incarcerated peers (log-rank **χ**^2^(1)=189.1, P<0.001) (**Figure 2B**). Strikingly, there was a decade difference between the groups in terms of median survival probability (69 vs. 79). That is, the age at which half of HRS participants were estimated to have experienced cognitive impairment came a decade earlier for the group with a history of incarceration compared to those without such history.

A Cox proportional hazard model confirmed that lifetime incarceration was associated with earlier cognitive impairment independent of covariates (HR=1.33, 95% CI [1.17-1.51], P<0.001) (**Table S1**, **Model S1.2**). When *APOE-ε4* genotype and lifetime incarceration were entered into the same model, their effect sizes did not change indicating independent risk pathways (**Model S1.3**). Similarly, the addition of multiplicative terms did not indicate the presence of unexplained excess risk for participants who experienced incarceration and had one or more *APOE-ε4* alleles (**Model S1.4**).

## Discussion

Cognitive decline is a hallmark of aging and a prerequisite for neurodegenerative diseases like Alzheimer’s disease. The progression toward cognitive impairment and eventually dementia can be exacerbated by factors both genetic and environmental. Here, we examined two such risk factors–one well-known (*APOE-ε4* genotype) and one less well-known (incarceration history)– and sought to determine the form of their interplay. We observed that *APOE-ε4* genotype and lifetime incarceration were independently related with an increased risk for cognitive impairment among HRS participants (i.e., a “G+E model” of risk).

Surprisingly, when estimating overall risk for cognitive impairment, the effect size for lifetime incarceration was identical to that for carrying two copies of the *APOE-ε4* allele, considered to be the strongest genetic risk factor for Alzheimer’s disease.(46) We found that people with a history of incarceration developed cognitive impairment a full decade earlier than people who do not have a history of incarceration. These findings position lifetime incarceration as an environmental risk factor for cognitive impairment that is comparable to *APOE-ε4* genotype, the leading genetic risk factor for Alzheimer’s disease.

The above comparison of risks/hazards between *APOE-ε4* genotype and lifetime incarceration should be interpreted with caution as *APOE-ε4* genotype is a single, precisely measured risk factor and past incarceration is likely a proxy (i.e., not causal) for the many exposures that go along with a criminal lifestyle. Nonetheless, we believe that criminal justice contact, particularly at the level of incarceration, is an important indicator to consider in population health research for three reasons. First, past incarceration identifies a segment of the population that is disproportionately poor, low in academic achievement, and hailing from minoritized groups(47–51) all risk factors for neurodegenerative diseases. Even if treated merely as an indicator of the concentration of risk, past incarceration would be valuable information for research aiming to characterize the health burden in a population. Second, there is evidence that past incarceration is not merely an indicator of the underlying risk of the (highly selected) carceral population, but also causally contributes to the health burden of inmates via direct (i.e., exposures during their sentence) or indirect means (i.e., incarceration-related exposures experienced after release), though this line of research is still nascent.(28, 52, 53) Third, incarceration may exacerbate underlying risks (e.g., genetic risk factors) in a multiplicative fashion, going beyond the mere additive effects. The current study did not find evidence for the latter possibility (i.e., no statistical interaction between past incarceration and *APOE-ε4* genotype was observed) but we believe that such a scenario is worth examining in the case of other outcomes, especially age-related conditions with different genetic profiles (e.g., polygenic instead of oligogenic).

The current study relied on participants in a population health survey. This approach is imperfect as it assumes that the population of interest (i.e., those affected by the incarceration experience) are likely to participate in population health surveys after their return to the general population. To understand the impact of incarceration on cognitive outcomes more fully (and the progression of age-related conditions more generally) we suggest that researchers move their attention to variation occurring during the incarceration experience itself. Our results comport with a litany of studies underscoring the necessity of risk assessment and classification for offenders during the intake process (i.e., right before they enter prison). Traditional assessments, such as the Level of Service Inventory-Revised (LSI-R), typically focus on indicators of criminogenic risk including antisocial cognitions and personality patterns and have been empirically validated over time and space for both males and females.(54–56) More recently, scholars have emphasized the importance of embedding indicators of psychiatric and medical afflictions into these tools as a means of improving service provision.(57, 58) There have also been calls to provide in-depth assessments and evaluations of the aging process within prisons (59) to many issues posed by members of the “graying” prison population and the “accelerated aging” that happens behind bars.

According to the United States Census Bureau, the threshold for elderly status among community samples is approximately 65 years old; by contrast, it is, on average, much lower among correctional populations and, according to the National Institute of Corrections, generally includes anyone over the age of 50.(60, 61) Scholarship indicates that incarcerated offenders over the age of 50 maintain health profiles equivalent to persons outside of prison who are 65 and older.(62) Given the results of the current study, administrators might consider specialized or alternative placements for individuals with higher levels of genetic risk on the *APOE-ε4* genotype as a means of anticipating some of the financial and logistical encumbrances (e.g., healthcare spending and facility layout/design) that accompany the management of elderly offenders.(63) In addition to identifying individuals based on genetic risk, prison officials may also consider administering a battery of validated cognitive tests, such as the Montreal Cognitive Assessment, to offenders during the intake process to determine baseline levels of functioning and permit tracking over time to inform future decisions (such as compassionate release). This is particularly important because manifestations of genetic risk do not remain static over time; rather, they can be curbed or exacerbated, depending on the prison experience and are crucial to the establishment of causal ordering/temporality when examining the incarceration-cognitive impairment nexus. Consider, for example, individuals suffering from traumatic brain injury (TBI). TBIs vary in their severity and include “mild,” “moderate,” and “severe” classifications; yet an individual who enters prison with a mild TBI could end up in the severe category if (s)he is exposed to any number of prison stressors, including violence. Of course, such considerations must be balanced against other indicators of risk, including the history of and propensity for violence(64), as well as the ability to maintain and provide safety and security to others.

Disentangling the incarceration-neurodegeneration nexus will become of critical importance in the coming decades, as both the number of individuals living with ADRDs in the community and the “graying” of the prison population are projected to increase substantially. For example, recent estimates indicate that approximately 6 million individuals in the U.S. are currently living with ADRDS; by 2060, this figure will have more than doubled and directly impact nearly 14 million people. By the same token, as of 2020, approximately 20% of the U.S. prison population met the age threshold to be considered “elderly” (≥50 years old); by 2030, this figure will rise to approximately 33%.(21, 48) The results of the current study and aforementioned projections comport with suggestions made recently by Testa *et al*(24) who emphasized the necessity of studying the nexus between incarceration and ADRDs throughout the life course because of its potential impact on (1) the reentry and reintegration of affected individuals, noting that it may increase social and economic disparities which independently correlate with general wellbeing; (2) the efficacy of treatment from caregivers and community service providers after an individual’s release; and (3) our understanding of the causal ordering associated with incarceration, ADRDs, and life expectancy (i.e., causation vs. selection and reverse causality).(65, 66) In their call for research, they emphasize the importance of analyzing high-quality data sources. In the current study, we heeded Testa and colleagues’ (2023) call by examining data from the Health and Retirement Study (HRS), with an explicit focus on the enduring effects of imprisonment (i.e., beyond the post-incarceration mortality spike).

The results and limitations of the current study offer several avenues for future research, including issues identified by Testa *et al*.(24) First, the HRS did not collect information regarding the timing of incarceration, which prevented us from establishing temporal order. The lack of temporal order precludes our ability to examine many relevant aspects of the association between incarceration and cognitive decline. For instance, it is impossible to establish whether a third variable, like drug use, acts as a mediator or a confounder. To this end, the data do not permit use to test for reverse causality, and specifically the extent to which cognitive reserves and impairment affect the probability of incarceration. As Testa *et al.* note, metrics of impulsivity, intelligence, and brain injury independently correlate with incarceration as well as cognitive decline. Future research should therefore consider these and other relevant factors when studying the incarceration-cognitive impairment nexus to parse out causal pathways.

Second, the measurement of lifetime incarceration in the HRS combines many different forms of incarceration into a single indicator, likely concealing a large degree of heterogeneity in the carceral experience. The incarceration experience (i.e., the “dose”)(24) varies across many dimensions including but not limited to the period, age of onset, duration, chronicity, security level, and subjective experience. It should therefore be a public health priority to understand which dimensions impart the greatest risk for cognitive decline, thus allowing more targeted policy conversations. Finally, participants of population health studies such as the HRS are not always representative specific sub-populations of interest to researchers (e.g., the formerly incarcerated population). It is possible that the current results were affected by this form of ascertainment bias, especially if those with incarceration experience and *APOE-ε4* risk alleles were disproportionately less likely to participate in the study. We do not believe this specific limitation to have greatly impacted our results, however, given that no difference in rates of *APOE-ε4* genotype between incarcerated/non-incarcerated groups were present in our analytic sample (**Table 1**).

## Conclusion

Incarceration is a heterogenous exposure with many downstream consequences for later life, some of which put former inmates at increased risk for cognitive decline. We found that incarceration imparted risk for cognitive impairment in later life that was independent of and comparable to the leading genetic risk factors. As a risk factor, past incarceration is potent but also opaque. More exploration of the mechanistic processes underlying the incarceration-cognitive impairment nexus may contribute to the amelioration of population-wide disparities in age-related conditions.

## Data Availability

Most of the data used in this study is publicly available from the HRS website: https://hrs.isr.umich.edu/. However, data on APOE genotypes are considered sensitive and require the submission of a sensitive data access use agreement.

## Supplemental material

**Table S1.** Cox proportional hazard model of lifetime incarceration and *APOE-ε4* genotype on first cognitive impairment.

| Variable<sup>1</sup> | Model S1.1 |  | Model S1.2 |  | Model S1.3 |  | Model S1.4 |  |
| --- | --- | --- | --- | --- | --- | --- | --- | --- |
|  | HR<sup>2,3</sup> | 95% CI<sup>3</sup> | HR<sup>2,3</sup> | 95% CI<sup>3</sup> | HR<sup>2,3</sup> | 95% CI<sup>3</sup> | HR<sup>2,3</sup> | 95% CI<sup>3</sup> |
| *APOE-ε4*<br>allele count |  |  |  |  |  |  |  |  |
| One copy | 1.19*** | [1.08, 1.31] |  |  | 1.19*** | [1.08, 1.31] | 1.20*** | [1.08, 1.33] |
| Two copies | 1.56*** | [1.20, 2.02] |  |  | 1.57*** | [1.21, 2.04] | 1.58** | [1.20, 2.08] |
| Lifetime<br>incarceration |  |  | 1.33*** | [1.17, 1.51] | 1.33*** | [1.17, 1.51] | 1.36*** | [1.17, 1.57] |
| Lifetime<br>Incarceration |  |  |  |  |  |  |  |  |
| × One copy |  |  |  |  |  |  | 0.94 | [0.72, 1.22] |
| × Two copies |  |  |  |  |  |  | 0.97 | [0.41, 2.29] |

<sup>1</sup> All models adjusted for sex, race/ethnicity, high school completion, smoking history, stroke history, and social origins index. All models were stratified by HRS cohort.
<sup>2</sup> \*p<0.05; \*\*p<0.01; \*\*\*p<0.001
<sup>3</sup> HR = Hazard Ratio, CI = Confidence Interval

**Figure S1.**
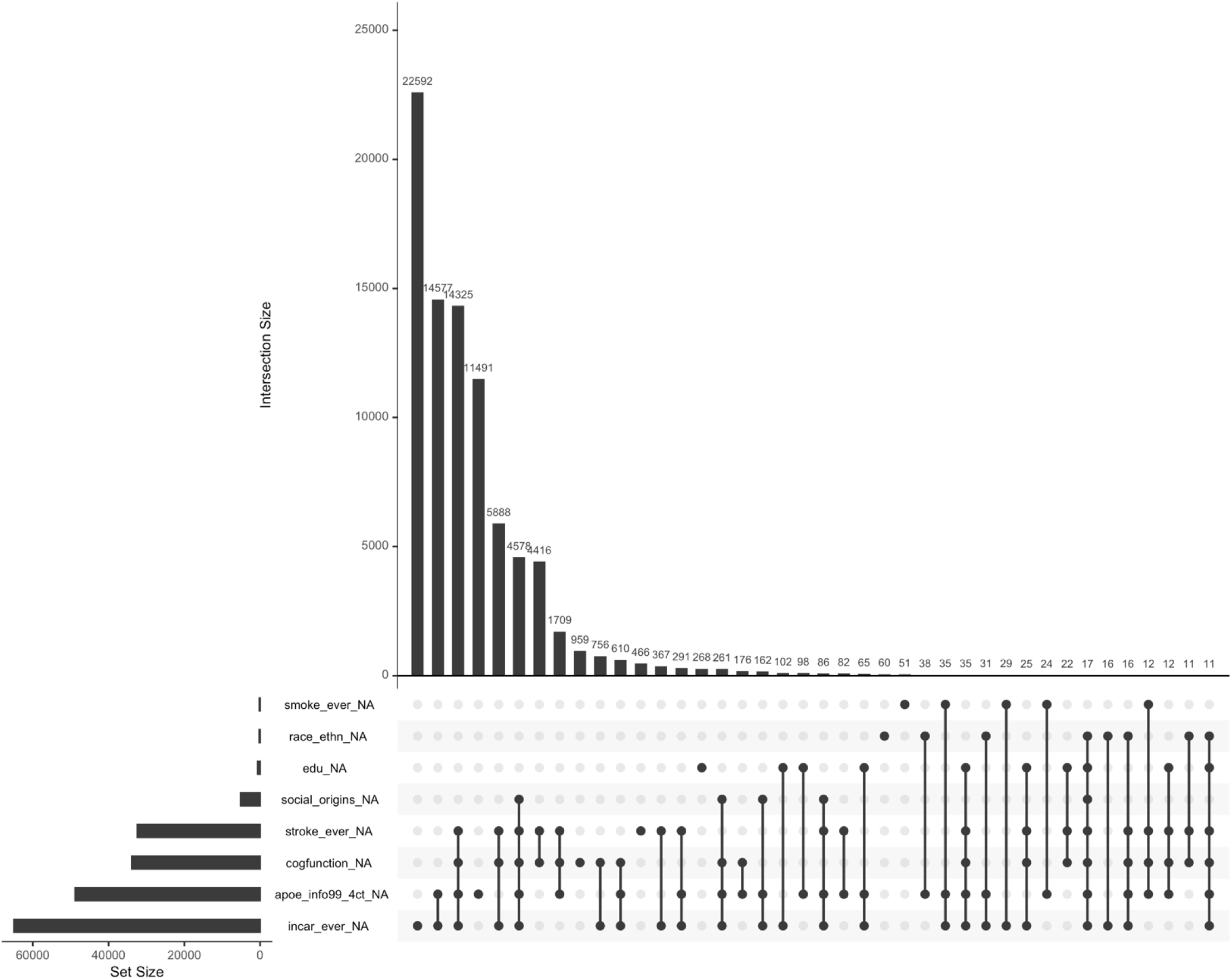
Patterns of missingness across study variables. Missing data counts refer to observations, not cases. Prior to listwise deletion, the sample included *N*=23,797 cases and *N*=141,392 observations. After restriction to complete cases, the analytic sample contained N=6,949 cases and N=55,345 observations. Note: only variables with missing values are displayed; HRS participants from the HRS and LBB cohorts were not included in the current analysis due to small case counts (5 for HRS) and recruitment after 2012/2014 (LBB) when incarceration variables were administered. Upset plot was created using the R package *naniar*.(67)

